# Centenarians and extremely old people living with frailty can elicit durable SARS-CoV-2 spike specific IgG antibodies with virus neutralization functions following virus infection

**DOI:** 10.1101/2021.03.05.21252707

**Authors:** Mary K. Foley, Samuel D. Searle, Ali Toloue, Ryan Booth, Alec Falkenham, Darryl Falzarano, Salvatore Rubino, Magen E. Francis, Mara McNeil, Christopher Richardson, Jason LeBlanc, Sharon Oldford, Volker Gerdts, Melissa K. Andrew, Shelly A. McNeil, Barry Clarke, Kenneth Rockwood, David J. Kelvin, Alyson A. Kelvin

## Abstract

**Background:** The SARS-CoV-2 (Severe Acute Respiratory Syndrome coronavirus 2) has led to more than 114 million COVID-19 cases and >2.5 million deaths worldwide. Epidemiological analysis has revealed that the risk of developing severe COVID-19 increases with age. Despite a disproportionate number of older individuals and long-term care facilities being affected by SARS-CoV-2 and COVID-19, very little is understood about the immune responses and development of humoral immunity in the extremely old person after SARS-CoV-2 infection. Here we investigated the development of humoral immunity in centenarians following a SARS-CoV-2 outbreak in a long-term care facility.

**Methods:** Extreme aged individuals and centenarians who were residents in a long-term care facility and infected with or exposed to SARS-CoV-2 were investigated for the development of antibodies to SARS-CoV-2. Blood samples were collected from positive and bystander individuals 30 and 60 days after original diagnosis of SARS-CoV-2 infection. Plasma was used to quantify IgG, IgA, and IgM isotypes and subsequent subclasses of antibodies specific for SARS-CoV-2 spike protein. The function of anti-spike was then assessed by virus neutralization assays against the native SARS-CoV-2 virus.

**Findings:** Fifteen long-term care residents were investigated for SARS-CoV-2 infection. All individuals had a Clinical Frailty scale score ≥5 and were of extreme older age or were centenarians. Six women with a median age of 98.8 years tested positive for SARS-CoV-2. Anti-spike IgG antibody titers were the highest titers observed in our cohort with all IgG positive individuals having virus neutralization ability. Additionally, 5 out of the 6 positive participants had a robust IgA anti-SARS-CoV-2 response. In all 5, antibodies were detected after 60 days from initial diagnosis.

**Interpretation:** Extreme older frail individuals and centenarians were able to elicit robust IgG and IgA antibodies directed toward SARS-CoV-2 spike protein. The antibodies were able to neutralize the virus. Humoral responses were still detectable after 60 days from initial diagnosis. Together, these data suggest that recovered participants who are of extreme old age would be protected if re-exposed to the same SARS-CoV-2 viral variant. Considering the threat of SARS-CoV-2 and COVID-19 to older age groups and long-term care facilities, the humoral responses to SARS-CoV-2 in older age groups is of public health importance and has implications to vaccine responses.

**Funding:** Canadian Institutes of Health Research (CIHR), NIH/NIAID, Genome Atlantic. VIDO receives operational funding from the Canada Foundation for Innovation through the Major Science Initiatives Fund and by Government of Saskatchewan through Innovation Saskatchewan.

## Introduction

In December 2019, a novel coronavirus was identified in Wuhan, Hubei China, which has led to a devastating pandemic (WHO pandemic)^1,2^. Coronavirus disease 2019 (COVID-19) is used to define the clinical symptoms that are associated with infection from severe acute respiratory syndrome coronavirus 2 (SARS-CoV-2)^3^. The SARS-CoV-2 is an enveloped positive-sense, single-stranded RNA genome and belongs to the *Coronaviridae* family in the genus *Betacoronavirus, sharing the* same lineage (Lineage B) as the 2002 SARS-CoV^4^. The spike protein of SARS-CoV-2 is an external binding protein, which guides the virus to attach to a host cell and bind the angiotensin converting enzyme II receptor (ACE2)^5^. The spike protein is the primary immunogenic target for virus neutralization and vaccine design, due to its critical role in the virus life cycle^5^.

As the clinical cases of SARS-CoV-2 infection were analyzed, it was clear that signs and symptoms, clinical manifestations, and disease severity widely varies ^6,7^. Certain host factors have been linked to the development of severe COVID-19 resulting in pneumonia, acute respiratory distress syndrome (ARDS), and multi-organ failure ^8,9^. In particular, age has been identified as a risk factor for severe illness and death with the elderly population being most susceptible^8^. It is suspected that age-related changes to the immune system, including immunosenescence and ‘inflamma-aging’ as well as the increased susceptibilty to co-morbidies, may contribute to increased risk of severe COVID-19 in older individuals ^10,11^. Additionally, those at the highest risk are older adults residing in long-term care homes^12,13^ due to the increased number of individuals living in one area^12^, asymptomatic transmission^14–16^, atypical symptom presentation in older people ^12,15^, and the high burden of chronic illnesses ^12,17^. Although older individuals are heavily represented in COVID-19 case fatalities, the clinical course and immune responses of older persons to SARS-CoV-2 infection is poorly understood ^11^.

Here, we examine a cohort of extreme old residents of a long-term care home that experienced a COVID-19 outbreak in Nova Scotia, Canada, with a focus on a group of centenarians who were infected with SARS-CoV-2. The humoral response and durability of antibody production following SARS-CoV-2 infection is currently ill-defined although multiple reports have documented a spectrum of responses in several cohorts. The role of age as a cofactor in establishing protection against future SARS-CoV-2 re-exposure and reinfection is of considerable interest in aged individuals. We investigated the antibody responses to SARS-CoV-2 infection in a small group of extreme aged individuals to better understand how the aged immune system works to protect against SARS-CoV-2 infection.

## Research in Context

### Evidence before this study

At this time, we are not aware of any reports on the seroconversion ability, durability, antibody function or antibody isotype landscape of centenarians infected with SARS-CoV-2. There are many studies in non-centenarians in which neutralizing antibodies have been detected in convalescent participant plasma, with primary focus on IgG and IgM titres post SARS-CoV-2 infection. It is well documented that long-term care facilities and older individuals are more susceptible to severe outcomes of SARS-CoV-2 infection and COVID-19, but little is understood regarding the older individual’s immune response to the virus during infection.

### Added Value of this Study

To our knowledge, this is the first study investigating seroconversion in SARS-CoV-2 infected centenarians residing in long-term care. Our study demonstrates that an aged immune system is still capable of mounting an antibody response to SARS-CoV-2 infection and that the antibodies elicited have virus neutralizing ability. Our data suggests that older and frail individuals, such as those in our study, have the capacity to be protected from a second infection with SARS-CoV-2.

### Implications of all the available evidence

The findings demonstrate that serconversion tests could be used to diagnose previous viral infection, outbreaks, and assess possible immune protection in an accessible and efficient manner. Serology analysis of older individuals displaying aberrant clinical COVID-19 could be used for case tracking and contact tracing in long-term cares home which are known to experience increased transmission Further, our data has implications for vaccine effectiveness in older individuals.

## Methods

### Study Design and Participants

Residents of a non-profit long-term care (LTC) home in Halifax, Nova Scotia, between April and June 2020 where a widespread outbreak of SARS-CoV-2 occurred, were recruited to the study. Residents of the facility had single or double rooms. All residents of the LTC facility were considered exposed to the virus due to the widespread nature of the outbreak within the facility. Consenting participants were enrolled, and testing was performed using routine practices. Briefly, a flocked nasopharyngeal (NP) swab was collected in 3 mL universal transport media (Copan Diagnostics Inc., Murrieta, CA), or a combined oropharynx and anterior nares (OP/Na) swab was collected using the Aptima Multitest swab in 2.9 mL of specimen transport medium (Hologic, Inc., San Diego CA). NP or OP/Na swabs were subjected to real-time RT-PCR using the SARS-CoV-2 assay on a Cobas 6800 system (Roche Diagnostics, Mississauga, Ontario, Canada) per the manufacturer’s instructions, or using a Total Nucleic Acid (TNA) extraction on a MagNApure LC 2.0 instrument (Roche Diagnostics) followed by real-time RT-PCR using a laboratory-developed test (LDT) designed at the British Columbia Centre for Disease Control (BCCDC) (Vancouver, BC)^18^. Twice a year, residents of Northwood each receive an update to their long-term care comprehensive Geriatric Assessment^19^. It includes a Clinical Frailty Scale score^20^. The CFS is a well-validated measure which categorizes frailty from fit to severely frail. Here we extracted information for analysis: age, sex, frailty, comorbidities, BMI (body mass index), and time between identification of the positive SARS-CoV-2 RT-RNA PCR test and blood collection^21^. A clinical description of the cohort is summarized in **Table 1**.

**Table.**
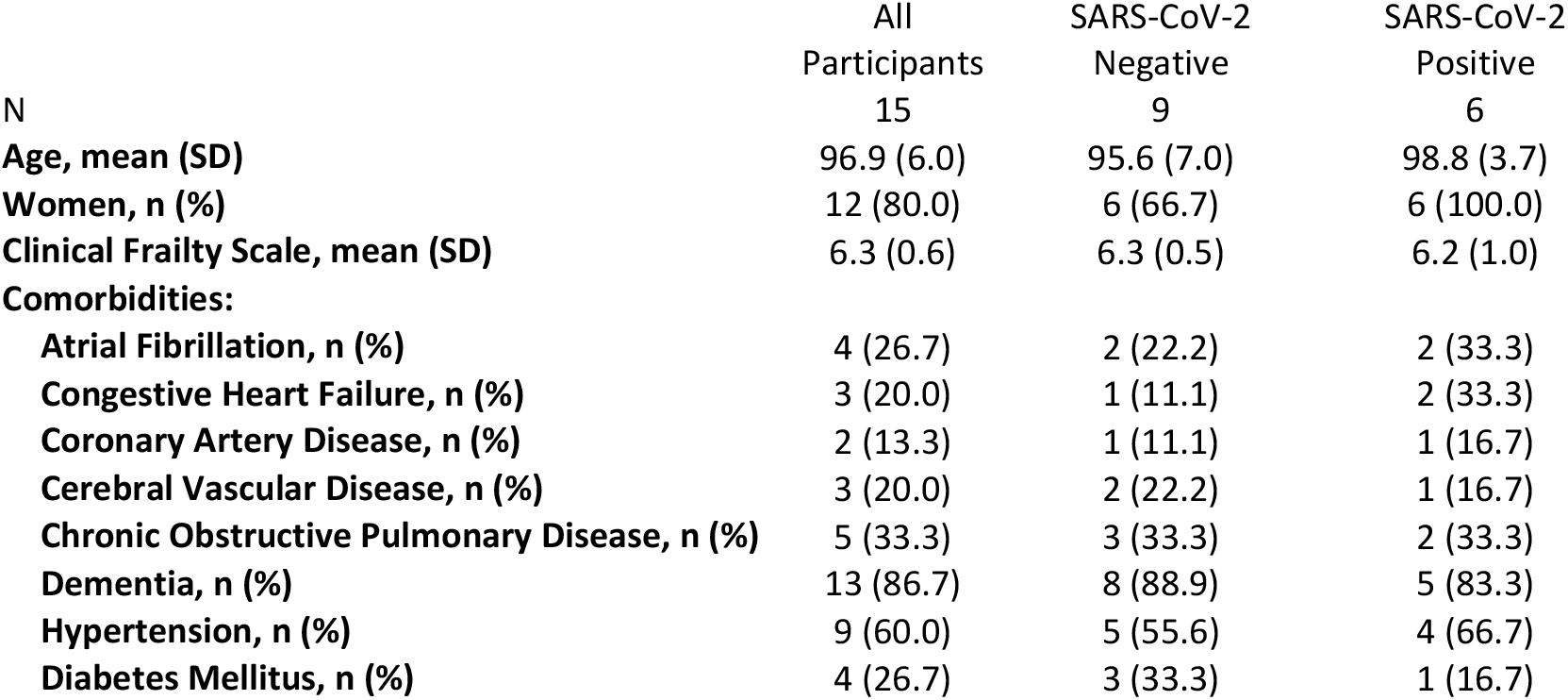
Clinical data from SARS-CoV-2 exposed and infected extreme aged residents of a long-term care facility.

### Blood Sample Processing

Peripheral blood was collected from study participants in K2EDTA spray coated tubes (Fisher 367861). Blood samples were immediately transported to the laboratory and centrifuged for 10 m at 200 x g after collection. Plasma was isolated and stored in conical vials (Fisher 1495949B) at −80°C until assays were performed.

### Spike Protein Synthesis

To acquire SARS-CoV-2 Spike (S) protein for molecular assays, a truncated S (NCBI data base reference sequence (NC_045512.2)) was cloned into pDSG-IBA 103 Expression Vector. The furin cleavage site was mutated and the cloned product was validated by DNA sequencing at Genewiz (South Plainfield, New Jersey, USA). For S protein production, Mexi293E cells were grown in 6 well cell culture plates. For transfection, 3 µg of plasmid was diluted in 150 µl serum-free OptiMEM. In a separate tube, 10 µl of Polyethyleneimine (PEI) (1mg/ml) was diluted in 150 µl Opti-MEM, vortexed briefly, and incubated for 5 m. The two solutions were combined, gently vortexed, and then incubated for 20 m. Culture media on the cells was replaced with DMEM + 5% FBS, and the PEI/DNA mixture was added. After incubation for 72 h at 37°C in a 5% CO2 chamber, recombinant S protein was harvested in cell culture media. S protein was purified using the IBA Strep-Tactin XT® Superflow® High Capacity resin system. Purified S protein was then used for ELISA assays.

### Enzyme-Linked Immunosorbent Assay

Plasma SARS-CoV-2 antibodies were detected using an indirect enzyme linked immunosorbent assay (ELISA). Microplates (96-well) (Corning^**®**^ 9018) were coated with 50 μl of 2.5 μg/mL SARS-CoV-2 S protein (in house or Sino Biology 40591-V08H) diluted in phosphate buffered saline (PBS) overnight at 4°C. Plates were washed three times with 1x PBS (Gibco 70011069) supplemented with 0.1% Tween-20 (Sigma P1379) (PBS-T). Plates were blocked with 5% BSA (Bovine Serum Albumin) in PBS-T for one h at room temperature. Blocking solution was removed, and 100 μl of human plasma samples diluted in 5% BSA PBS-T were diluted across plates and incubated for two hours at room temperature. Plates were washed three times with PBS-T. Secondary antibody was diluted in 1% BSA in PBS-T as follows: anti-human IgG-HRP 1:2000 (Invitrogen 31413), anti-human IgM-HRP 1:2000 (Invitrogen A18835), anti-human IgA-HRP (Invitrogen A18781). Plates were incubated in secondary antibody for one h at room temperature. Plates were then washed five times with PBS-T, and 50 μl TMB substrate (Thermo 34022) was added to each well. After 15 m, 50 μl of stop solution (Invitrogen SS04) was added to each well and plates were read at 450nm on a plate reader (Bio Tek Synergy LX Multi-Mode Reader (<u>BTSLXFATS</u>). Samples were considered positive if average optical density (OD) was greater than 0.1 and greater than the mean OD in SARS-CoV-2 unexposed samples plus 3 standard deviations at the same dilution. Negative samples are denoted as “1” for display on a logarithmic scale.

### SARS-CoV-2 Virus and Culture

The SARS-CoV-2 strain SARS-CoV-2/Canada/ON/VIDO-01-2020 was used for *in vitro* neutralization assays. The virus was isolated from the respiratory secretions from an infected 56-year-old man presenting with respiratory symptoms at a hospital in Toronto, Canada, upon returning from Wuhan, China ^22^. The viral stock used was from the second passage and the sequence is available at GISAID – EPI_ISL_425177. The virus was amplified in Vero-76 cells in viral growth media vDMEM (Dulbecco’s Modified Eagle Medium (*Wisent Bioproducts (Cat # 319-005-CL)*), 2% fetal calf serum (*Wisent Bioproducts (Cat # 090-150)*), 5 mL 100x Penicillin (10,000 U/mL)/Streptomycin (10,000 ug/mL), and 2 mg/mL TPCK-trypsin) at VIDO, Saskatoon, Saskatchewan. All work with SARS-CoV-2 live virus was performed in a CL3 facility at VIDO.

### Neutralization Assay

Vero cells (ATCC® CRL-1587) were seeded into 96-well tissue culture plates (Corning) at 20,000 cells per well and incubated overnight at 37°C in a 5% CO2 chamber. Plasma samples were heat inactivated at 56°C for 30 m. Heat inactivated plasma was diluted 1:10 in serum-free DMEM and diluted 1:2 across round bottom 96-well plates (Corning). SARS-CoV-2 virus (SARS-CoV-2 strain /Canada/ON/VIDO-01-2020) was diluted to 100 TCID50/17.5μl in serum-free DMEM. The diluted virus was added to diluted plasma samples and incubated at 37°C for one hour. A back titer of the virus was also prepared to confirm the titer of the virus dilution used. The plasma-virus mixture was added to plated Vero 76 cells and incubated at 37°C in a 5% CO2 chamber for 1 h for virus absorption. The mixture was removed and replaced with vDMEM and placed back at 37°C with 5% CO2 for 3 days. Cells were checked daily for signs of cytopathic effect (CPE), and results were recorded on day 5 post inoculation. Antibody titer was calculated as the inverse of the most diluted sample where no CPE was detected. All work with SARS-CoV-2 live virus was performed in a CL3 facility at VIDO.

### Statistical Analysis

Results were analysed using GraphPad Prism8.

## Ethics

Ethics for this study was approved by the IRB at Dalhousie University and is covered under the protocol “Sentinel surveillance for severe outcomes of laboratory-confirmed influenza in adults for the annual influenza season and for confirmed and suspected cases of COVID-19/SARS-CoV-2 acute respiratory disease” REB#1020727.

## Results

A total of 15 SARS-CoV-2 exposed residents of a non-profit long-term care (LTC) facility were included. Their mean age was 96.9 (range 84 – 103). All residents of the LTC facility were considered exposed as their rooms were on floors with positive participants. All residents including these participants (12 women and 3 men) were routinely screened for SARS-CoV-2 infection by nasopharyngeal swabbing and subsequent qRT-PCR for the RNA encoding the SARS-CoV-2 nucleoprotein (NP) (**Table 1**). Six of the participants (all women) tested positive with a median age of the SARS-CoV-2 PCR positive group being 98.8 years (range 94-102). The median age of the SARS-CoV-2 PCR negative group was 95.6 years (range 84-103). Three of the SARS-CoV-2 NP PCR positive residents were centenarians, with ages of 100, 102, and 102 years. In total, there were 8 centenarians examined in the cohort. There were no differences in the level of frailty (Clinical Frailty Scale of 6.2 versus 6.3)^20^, or comorbidities (dementia, hypertension, chronic obstructive pulmonary disease, etc.) in those who were SARS-CoV-2 positive and SARS-CoV-2 negative, respectively (Table 1). As expected, there was a high burden of dementia and frailty in the study population. For plasma sample collection, the mean time from PCR diagnosis with COVID-19 to sample collection was 30 days. For residents with a second blood sample collected, the mean time between the first and second blood collection was 30 days.

An enzyme-linked immunosorbent assay (ELISA) was used to detect the presence of antibodies directed toward the SARS-CoV-2 S protein present in the first blood collection for each resident enrolled in the study. Subsequently, the isotype and subclasses of S antibodies were identified by ELISA as well. SARS-CoV-2 infected centenarians had high titres of S-directed IgG in their plasma, with endpoint titers ranging from 1:6400-1:52400 (**Figure 1Ai** and **C**). We were not able to detect S-directed IgG in the plasma of SARS-CoV-2 NP PCR negative participants (**Figure 1Aii** and **C**). Non-centenarians who tested PCR positive also had significant anti-S IgG endpoint titers (1:800) (**Figure 1Bi** and **C**). Antibody IgG subclass analysis indicated that both IgG1 and IgG3 S-directed antibodies were present in centenarian residents 862 and 916 and the non-centenarian residents (**Supplemental Figure 1** and **2**). Further isotype characterization revealed that all SARS-CoV-2 PCR positive centenarians had a detectable anti-S IgM response, with titres ranging from 1:400-1:6400 (**Figure 2Ai**) similar to positive non-centenarians with the exception of one positive non-centenarian who did not have detectable anti-S IgM (**Figure 2Aii**) summarized in **Figure 2C**. To complete the isotype analysis, S directed IgA antibodies were also measured in plasma samples where IgA1 anti-S levels were greater than IgA2 (**Supplemental Figure 2** and **3**). The SARS-CoV-2 positive centenarians were also positive for S IgA (1:800-1:25600) (**Figure 3**). To determine if the humoral responses to the SARS-CoV-2 virus were similar in magnitude between infected centenarian and non-centenarian older residents, we directly compared if the titres of S directed IgG, IgM, and IgA of the two groups. The titers of S directed IgG, IgM, and IgA were not statistically different between the groups, demonstrating that advanced age did not alter S-directed antibody elicitation to the SARS-CoV-2 virus.

**Figure 1.**
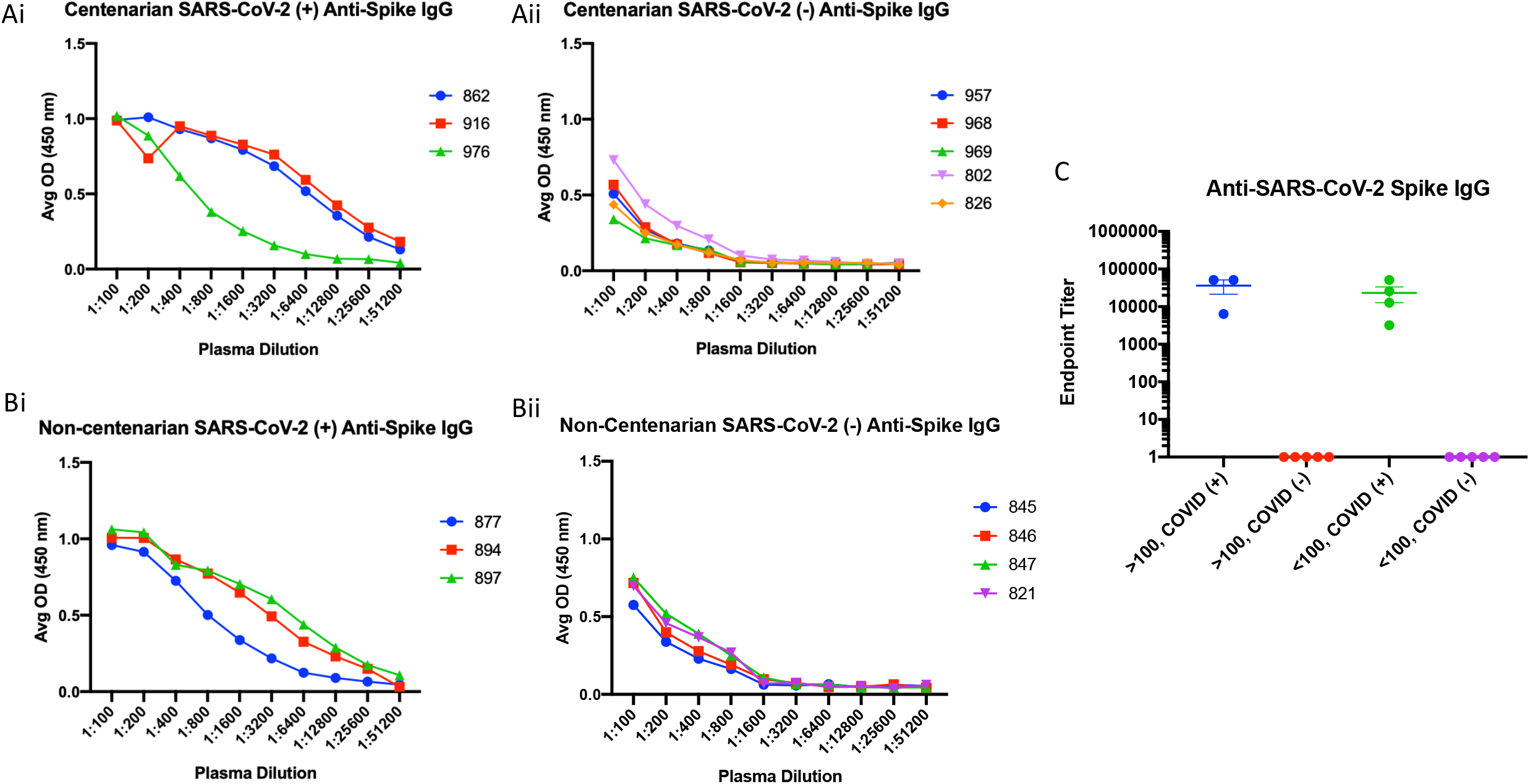
Extreme aged centenarians and non-centenarians infected with SARS-CoV-2 elicit a robust anti-spike IgG response. Peripheral blood samples were collected from 16 residents of a long-term care facility in Halifax, Nova Scotia, who were considered exposed to the SARS-CoV-2 virus. All exposed residents were tested for the presence of virus in the upper respiratory tract by nasopharyngeal swabbing following by PCR to detect the viral N gene. Blood was collected approximately 30 days after a positive PCR test. Serum separated was subjected to a direct ELISA assay to determine the magnitude of SARS-CoV-2 spike protein directed antibodies of the IgG isotype. The ELISA was analyzed by participant group: centenarians PCR positive for SARS-CoV-2 nucleic acid encoding gene N (**Ai**); centenarians PCR negative for SARS-CoV-2 nucleic acid encoding gene N (**Aii**); non-centenarians PCR positive for SARS-CoV-2 nucleic acid encoding gene N (**Bi**); non-centenarians PCR negative for SARS-CoV-2 nucleic acid encoding gene N (**Bii**). Endpoint titers for each group are summarized (**C**).

**Figure 2.**
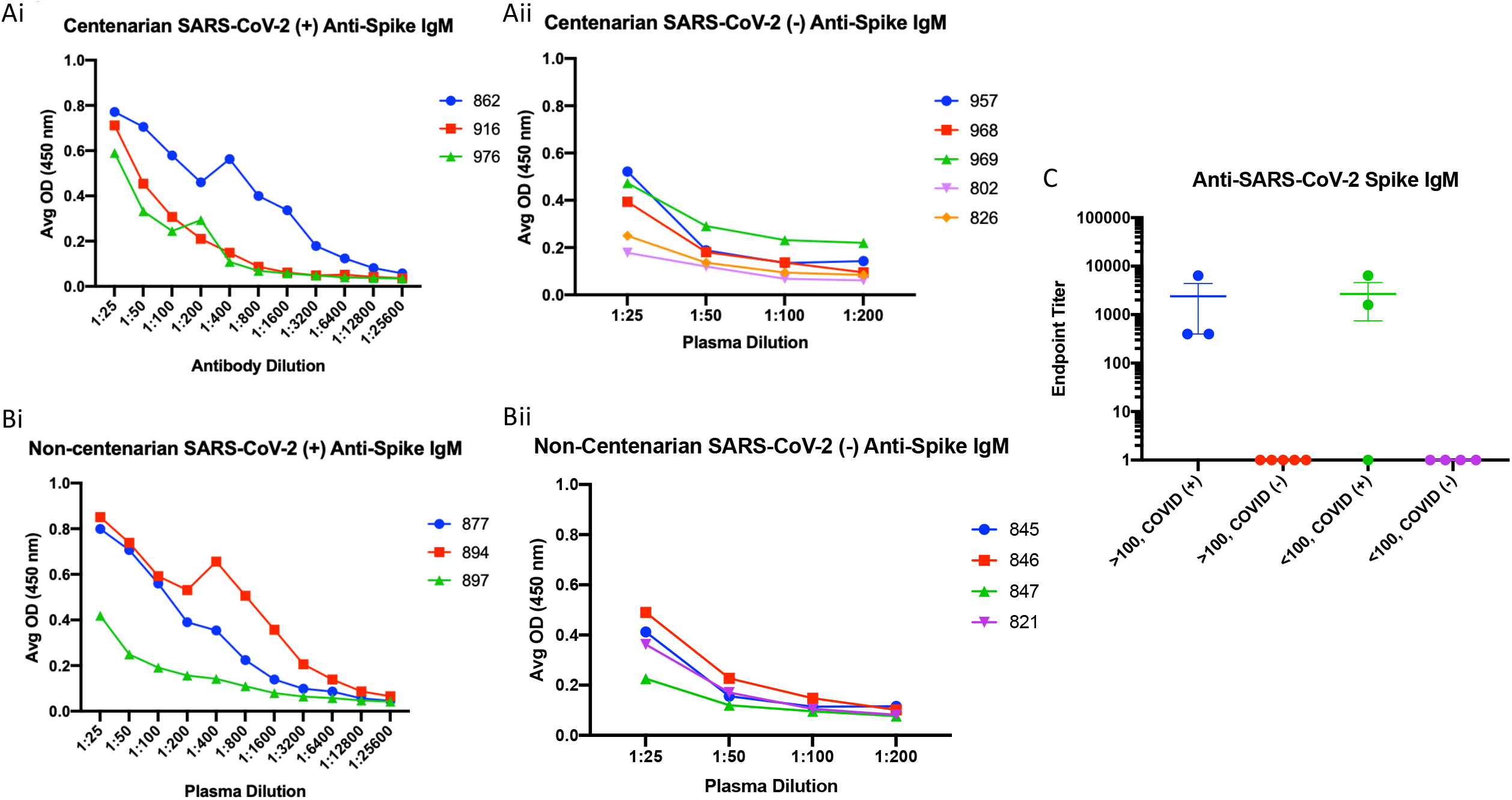
Similar profiles of spike IgM antibodies elicited in centenarians and non-centenarians with confirmed SARS-CCoV-2 infection. Plasma samples collected from the long-term care cohort were analyzed by participant group for the presence of anti-S IgM. The endpoint titer results were plotted by group: centenarians PCR positive for SARS-CoV-2 nucleic acid encoding gene N (**Ai**); centenarians PCR negative for SARS-CoV-2 nucleic acid encoding gene N (**Aii**); non-centenarians PCR positive for SARS-CoV-2 nucleic acid encoding gene N (**Bi**); non-centenarians PCR negative for SARS-CoV-2 nucleic acid encoding gene N (**Bii**). Endpoint titers for each group are summarized and compared (**C**).

**Figure 3.**
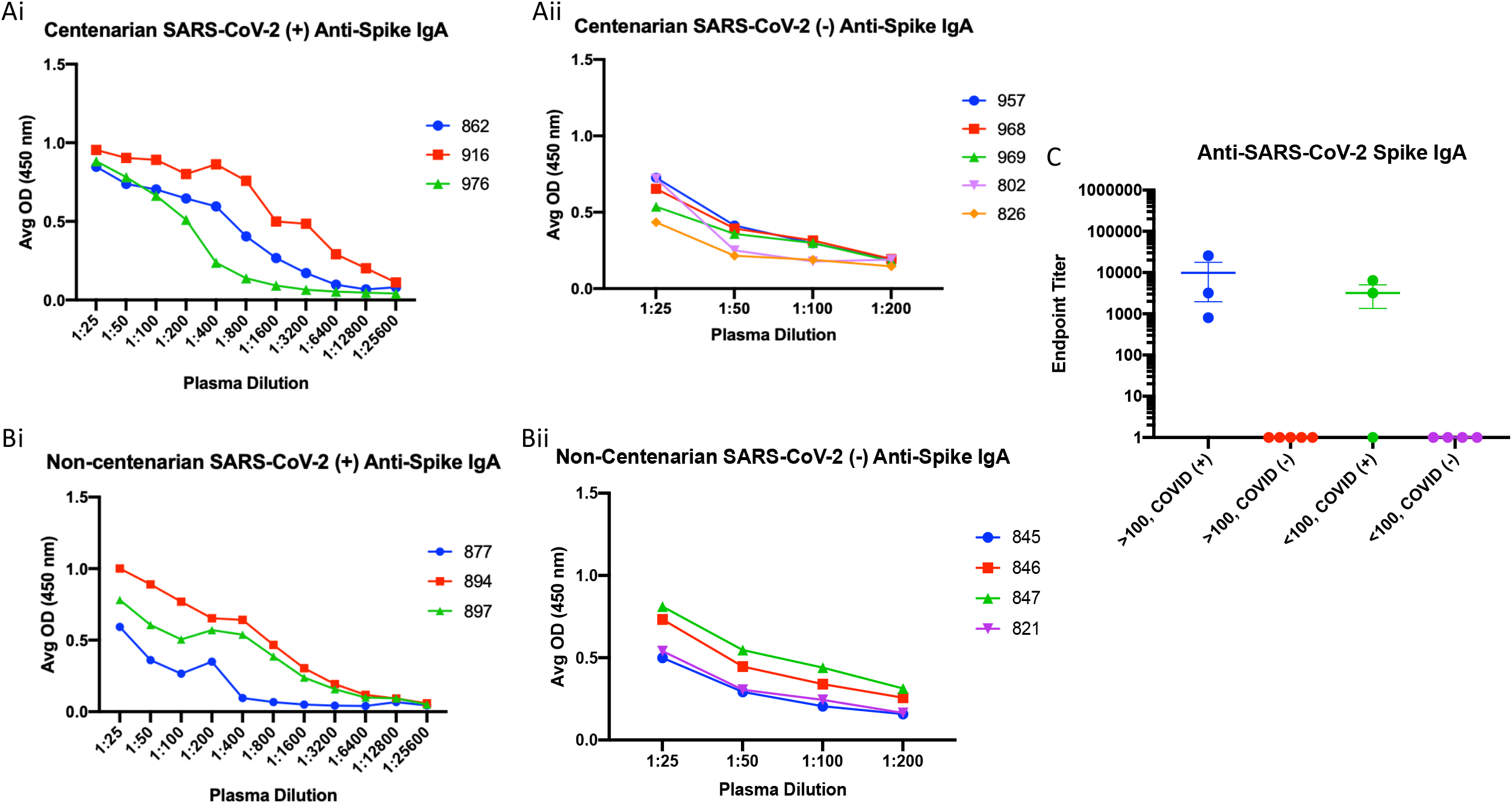
Anti-spike IgA is present in the plasma of centenarians and non-centenarians infected with SARS-CoV-2. Anti-S IgA was assessed in plasma samples collected from SARS-CoV-2 positive centenarians (**Ai**); SARS-CoV-2 negative centenarians (**Aii**); SARS-CoV-2 positive non-centenarians (**Bi**); SARS-CoV-2 negative non-centenarians (**Bii**). Endpoint titers for each group are summarized (**C**).

To investigate the durability and function of antibodies elicited toward SARS-CoV-2, we analyzed a second blood sample collected approximately 30 days after the first draw to determine levels of S antibodies and whether antibodies had viral neutralization capacity. For the durability assay, resident 916 had consistently high levels of anti-S IgG titres with levels of 1:52400 still observed in their second plasma sample (**Figure 4A**). Conversely, the titer of resident 976 decreased from 1:6400 to 1:3200 at the second draw. When evaluating anti-spike IgM, both residents had decreased responses (**Figure 4B**). Initial titers for both centenarians were 1:400 and decreased in the follow-up sample, with the titer in resident 916 IgM decreasing to 1:200. There was no detectable IgM for resident 976 at either time point. Decreases in antibody titer were also observed at the second timepoint for spike-directed IgA (**Figure 4C**). SARS-CoV-2 virus neutralization assays were performed to assess the functional neutralization capabilities of the plasma antibodies elicited by the COVID-19 advanced aged cohort. The plasma samples collected at the first sampling time point were used in standard virus neutralization assays performed in Vero76 cells with SARS-CoV-2 (strain SARS-CoV-2/Canada/ON/VIDO-01-2020) at 10 TCID50. All of the SARS-CoV-2 NP PCR and S IgG positive residents had detectable neutralizing antibody titers, ranging from 1:20 to 1:160 for centenarians and 1:20 to 1:40 for non-centenarians (**Figure 5**). Together, our data illustrates that centenarians infected with SARS-CoV-2 were able to elicit SARS-CoV-2 S-directed neutralizing antibodies, demonstrating an intact humoral immune response.

**Figure 4.**
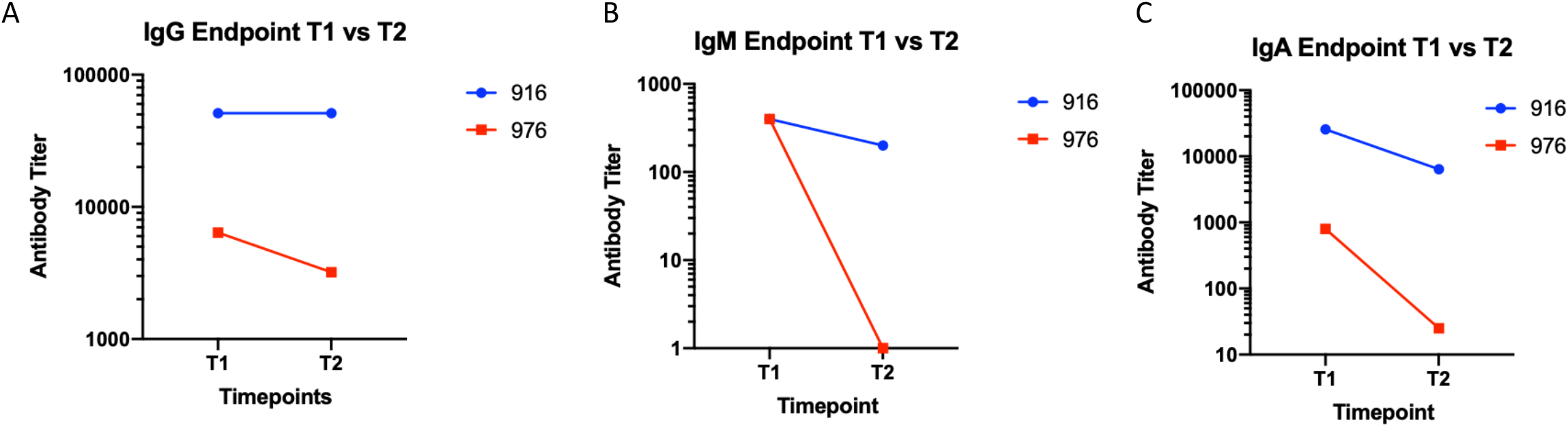
Anti-spike IgG remained robust 60 days after initial SARS-CoV-2 in centenarians. Anti-S IgG (**A**), IgM (**B**), and IgA (**A**) antibodies present in residents 916 and 976 plasma samples collected at 30 days (T1) and 60 days (T2) post a SARS-CoV-2 PCR test were assessed by ELISA.

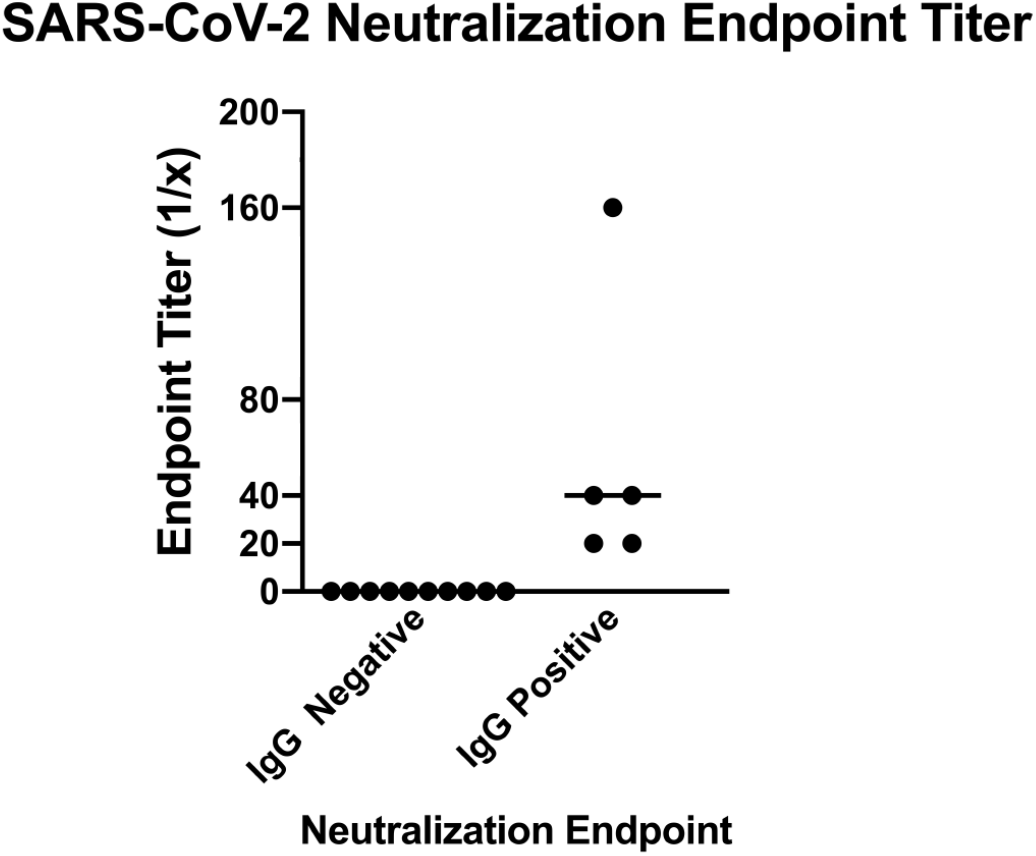

## Discussion

To our knowledge, this is the first study investigating seroconversion in SARS-CoV-2 infected extreme aged adults, some over 100 years of age, residing in a long-term care facility. Here, a group of highly aged, frail residents, the majority being female, residing in a long-term care facility who were infected with SARS-CoV-2 and survived, is described. The centenarians were able to elicit a successful S directed antibody response which was able to functionally neutralize the native SARS-CoV-2 virus. This work demonstrates that the extreme aged immune system is capable of responses during SARS-CoV-2 virus infection which are classically associated with convalescence and recovery from acute respiratory virus infection.

The older residents examined in this study represent a particularly vulnerable group in the ongoing global SARS-CoV-2 pandemic. Estimations of infection fatality risk (IFR) of SARS-CoV-2 in Switzerland demonstrated that people older than 65 years had an IFR of 5.6%, compared to 0.0092% in those aged 20-49 y ^23^. Moreover, risk of death from COVID-19 has been estimated to be 630 times greater for those over 85 y compared to 19-29 y/o ^24^. Older adults, especially those living with frailty, are considered disproportionately susceptible to COVID-19 as they have increased likelihood of chronic health issues and immune dysfunction^13,20^. Aged immune systems are characterized not only by immunosenescence, where the immune system deteriorates with time^12^, but also inflammaging, chronic, low-grade inflammation which can contribute to disease pathogenesis^10^. The weakened immune system in older adults has led to concern with their ability to mount a successful humoral immune response to SARS-CoV-2 infection considering their increased susceptibility to the virus.

Here, we showed that after an average of 30 days after a positive SARS-CoV-2 NP PCR test, some extremely old LTC residents mounted high SARS-CoV-2 S-directed antibody responses across relevant immunoglobulin isotypes. The highest titers were observed with IgG, where centenarian titres ranged from 1:6400 to 1:52400, which is associated with neutralization function 30 days post symptom onset at titers ranging from 1:20 to 1:160. Other studies have established ELISA IgG titres of 1:320 as moderate and titres of 1:960 and above as high ^25^, demonstrating that the centenarians in this study induced a robust anti-spike IgG response after SARS-CoV-2 infection. Interestingly, one study by Klein and colleagues indicated that serum SARS-CoV-2 antibody titers increased with age and male sex in COVID-19 patients ^26^. Since most of our advanced aged participants were female, our data suggests that extreme aged females also elicit high SARS-CoV-2 antibody titers. As well, our data demonstrate that even the most vulnerable members of the oldest old can mount an immune response and survive SARS-CoV-2 infection. In younger COVID-19 cohorts, by four weeks post symptom onset, over 90% of participants had neutralizing activity; the older residents in our study appear to have had a similar humoral response ^27^. Previous reports also indicate that S-directed IgG antibody titers have the strongest correlation with neutralization antibody titer in younger people infected with SARS-CoV-2 ^27^, agreeing with our results showing centenarians with anti-S IgG titers were also able to neutralize the virus.

The identified S-directed IgG response was durable and remained high at a second sample collection which was approximately 60 days after an initial diagnostic positive SARS-CoV-2 NP PCR test. Despite media concerns surrounding longevity of the humoral response for SARS-CoV-2 infection, our data, although collected from a small study group, suggest that S IgG antibody levels are slightly decreased 2 months after active infection, even in some extremely old people. A recent study investigating the durability of anti-S IgG has shown peak S-directed IgG antibodies 4 weeks after COVID-19 symptom onset which then remained high for 6 months ^27^. These results are in agreement with other North American studies that show stable SARS-CoV-2 S antibody levels to 115 days and 90 days post symptom onset (PSO) ^25,28^.

In contrast to our IgG anti-S findings, we found a robust early increase in S-specific plasma IgA in 5/6 SARS-CoV-2 infected people studied at 30 days post positive PCR test which declined. Few studies have characterized the onset of IgA after SARS-CoV-2 infection in plasma, most likely due to the mucosal association typically considered for the IgA isotype. One study that examined COVID-19 convalescent plasma found a correlation of S-specific IgG with S-specific IgA in plasma ^26^, as did we. Also, in agreement with our findings, another study found that plasma anti-S IgA peaked early (16-30 days post symptom onset) and declined to 74.1% of maximum titres by day 115 PSO ^28^. It remains unclear at this time how S-directed IgA antibody dynamics may influence COVID-19 disease severity and recovery from SARS-CoV-2 infection among all age groups.

Older people residing in long-term care homes represent an especially vulnerable population. Spread and transmission of the virus within long-term care has been shown to be very rapid ^15^. Within long-term care facilities in both Belgium and the USA, high proportions of asymptomatic cases were observed, leading to challenges with isolation of positive individuals within facilities^13,14,16^. Older people, especially those who are frail and have dementia, often develop subtle and atypical illness presentations that present challenges for symptom detection and lead to difficulty in identifying infection^29^. Within the cohort studied here, all but two residents were living with dementia and most residents were at least moderately frail. This may have contributed to the rapid spread of the virus through the long-term care facility. Frequent serological investigation and PCR testing for SARS-CoV-2 antibodies and virus, respectively, provide mechanisms for identifying an outbreak in extreme older individuals who may be unaware of their health status which is confounded by an atypical COVID-19 clinical picture. Another concern surrounding the immune response to SARS-CoV-2 in adults is the concept of cross-reactivity to the virus due to infection with other coronaviruses across the life span. Studies using highly-sensitive flow-cytometry based methods have shown that people who were not exposed to SARS-CoV-2 had detectable IgG antibodies against the spike protein of virus. These antibodies were especially prevalent in children and adolescents ^30^. Studies have also shown that unexposed people have pre-existing CD4+ memory T cells that are reactive to common cold coronaviruses such as NL63, HKU1, and OC43, as well as the current pandemic coronavirus SARS-CoV-2 ^31^. As the participants studied in this cohort were highly aged and had likely been exposed to many viruses across their lifespans, it was hypothesized that some cross-reactivity to the SARS-CoV-2 spike protein or virus may be observed. Interestingly, in our study, residents who did not test positive for SARS-CoV-2 RNA did not have any detectable plasma IgG against the SARS-CoV-2 spike protein. Our method of detection using indirect ELISA for IgG may suggest that cross-reactive antibodies from previous circulating common cold coronaviruses may not be an issue when screening individuals for serological evidence of a SARS-CoV-2 infection.

Our study was limited by a small sample size, lack of continued plasma sampling, and uncertain SARS-CoV-2 inoculation/exposure date. The low sample size reflects a low incidence rate in Nova Scotia, even among extremely old people living with frailty (cumulative confirmed cases were 177 per million on April 1 and 1057 by May 15, the period of initial data collection)^32^. Even so, the information gathered here is extremely valuable because of the rare occurrence of centenarians infected with SARS-CoV-2. The second timepoint for blood collection was not available for all residents studied, making it difficult to form more robust and broad conclusions from our data. Although more frequent blood collection would have been beneficial to better model the dynamics of the antibody response following infection in highly aged people, access to blood samples in this age group will remain a difficult task due to their vulnerability and the pragmatics of venipuncture. Finally, as with most viral infections, pinpointing the exact inoculation date or event for an infection is difficult in people. With increased studies following contact tracing and symptom onset as well as studies such as ours describing time from symptom onset and development of SARS-CoV-2 antibodies, a clearer clinical picture of disease progression and immune responses for extreme older adults as well as other age groups will be better defined.

The current COVID-19 pandemic has highlighted the often marked frailty of residents in long-term care which is evident by the high COVID-19 case numbers, COVID-19 fatalities, and SARS-CoV-2 outbreaks in these facilities across several countries ^33–37^. The cause of this may be multifaceted, which may include the inadequacies of infection control measures and gaps in education as well as the difficulty identifying atypical clinical manifestations of COVID-19 in older people. Our study shows that even the oldest people can elicit a strong humoral response to SARS-CoV-2 and recover from infection. These findings are important for developing serological testing protocols as well as investigating the poor COVID-19 outcomes associated with LTC facilities.

## Data Availability

All data can be accessed in the manuscript or raw values of ELISA assays can be requested if needed.

## Acknowledgements

We acknowledge the gracious help by Dr. Lisa Barrett with the study design, cohort management, and sample acquisition. We wish to thank all of the study participants for their involvement. This article is published with the permission of the Director of VIDO-InterVac with article number 932.

## Figure Legends

**Supplemental Figure 1.**
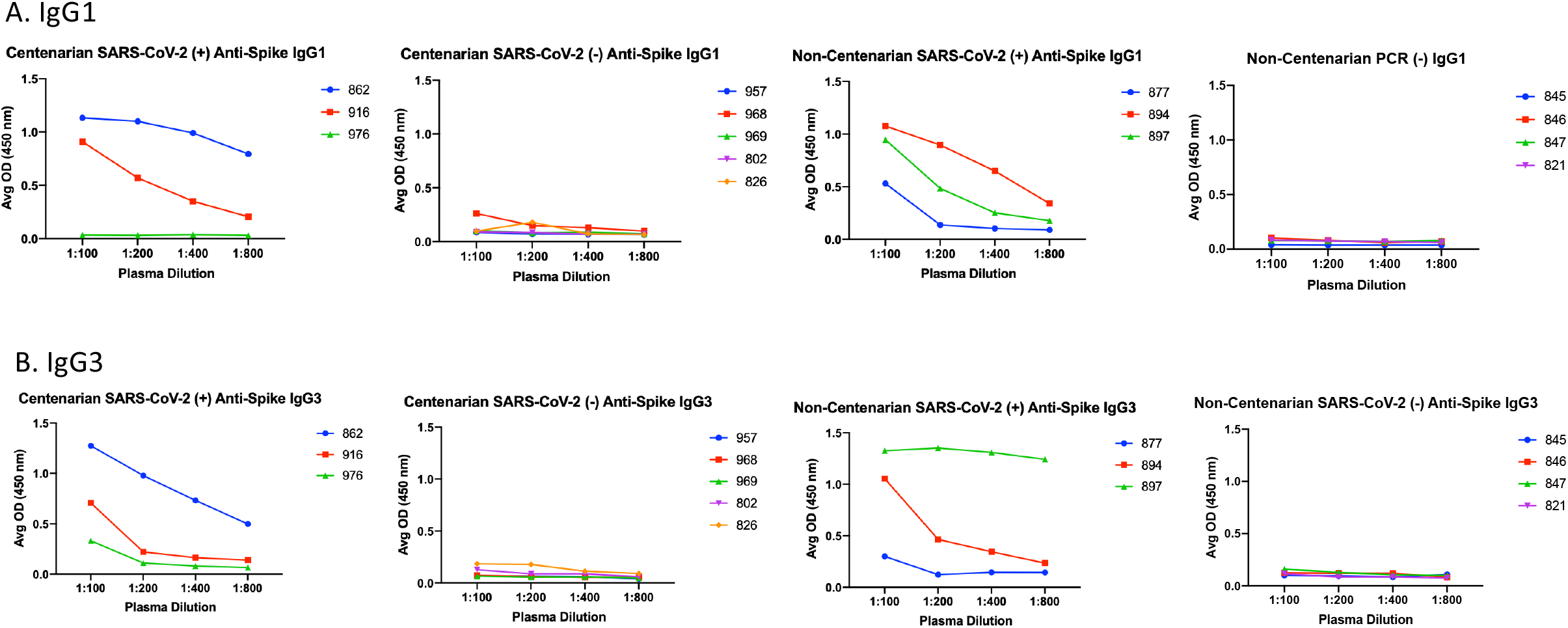
Extreme aged centenarians and non-centenarians infected with SARS-CoV-2 elicit IgG antibodies of IgG1 and/or IgG3 subclass. Plasma samples collected from long-term care residents 30 days after initial SARS-CoV-2 positive PCR tests or virus exposure were analyzed for IgG subclasses IgG1 (A) and IgG3 (B).

**Supplemental Figure 2.**
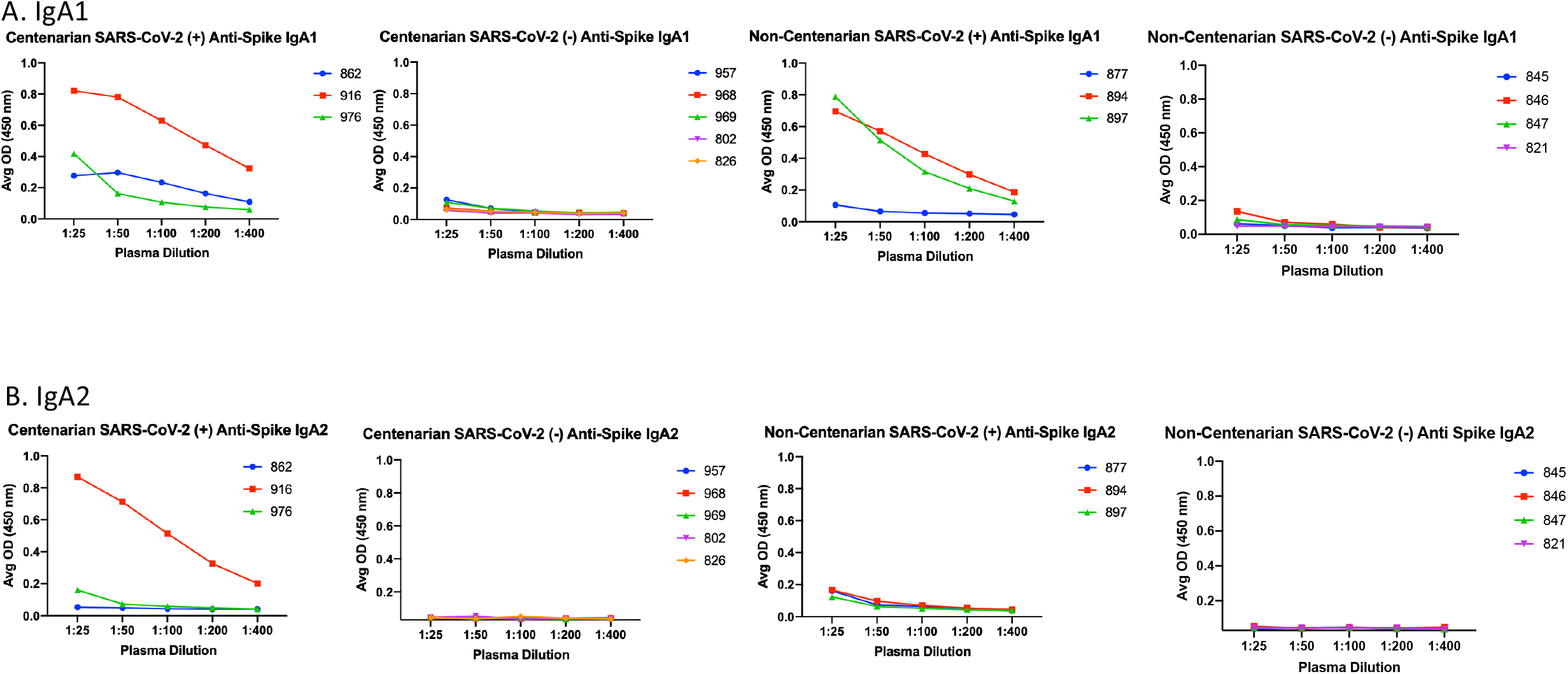
IgA subclasses elicited by SARS-CoV-2 extreme aged centenarians and non-centenarians are IgA1 skewed and/or IgA2 subclass. Plasma samples collected from long-term care residents 30 days after initial SARS-CoV-2 positive PCR tests or virus exposure were analyzed for IgG subclasses IgG1 (A) and IgG3 (B).

